# Pandemic-related decline in injuries related to women wearing high-heeled shoes: Analysis of U.S. data for 2016-2020

**DOI:** 10.1101/2021.12.26.21268426

**Authors:** Philip N. Cohen

## Abstract

**Background:** Wearing high-heeled shoes is associated with injury risk. During the COVID-19 pandemic, changes in work and social behavior may have reduced women’s use of such footwear.

**Methods:** This study assessed the trend in high-heel related injuries among U.S. women, using 2016-2020 data from the U.S. Consumer Product Safety Commission’s National Electronic Injury Surveillance System (NEISS).

**Results:** In 2020 there were an estimated 6,290 high-heel related emergency department visits involving women ages 15-69, down from 16,000 per year in 2016-2019. The 2020 decline began after the start of the COVID-19 shutdowns on March 15. There was no significant change in the percentage of fractures or hospital admissions.

**Conclusions:** The COVID-19 pandemic was associated with a decline in reported injuries related to high-heeled shoes among US women. If this resulted from fewer women wearing such shoes, and such habits influence future behavior, the result may be fewer injuries in the future.

## Introduction

Epidemiological studies find an association between wearing high-heeled shoes and hallux valgus, musculoskeletal pain, and osteoarthritis, which is supported by biomechanical research [1–3]. Beyond long-term harms, high-heeled shoe related injuries (HHSRIs) resulting in emergency department (ED) visits were reported for more than 14,000 U.S. women in 2012, with an increasing trend at least from 2002 to 2011 [4]. Women are much more likely than men to wear shoes that cause pain or discomfort – principally from high heels – and many acknowledge a tradeoff between such harms and their perceived benefits to attractiveness [5]. However, high heels may have begun losing market share in the last decade, and in 2019 the fashion press reported that runways models were wearing sneakers for women [6]. There was subsequent speculation that “2019 killed the heel” [7].

In this context, during the first year of the COVID-19 pandemic in the United States, the geographic mobility of much of the population was dramatically limited by both policy restrictions and their economic effects, and the independent decisions by many people to limit their movements in light of risks from social interaction [8–10]. In addition, in the second half of 2020, 26% of employed people worked from home at least some of the time because of the pandemic [11]. Reductions in mobility, especially resulting from less active socializing and more working from home, may have resulted in women wearing high heels less, leading to a reduction HHSRIs. However, it is also possible that women with less severe injuries stayed away from EDs during the pandemic, resulting in a decline in the percentage of injuries reported. If that is the case, we might expect to find fewer, but more severely injured, patients during the pandemic period.

## Materials and methods

The study used the Consumer Product Safety Commission’s National Electronic Injury Surveillance System (NEISS), which documents reports of ED visits at a sample of hospitals. Each visit is coded with up to 3 product codes. The analysis included incidents in which there was any use of code 1615, Footwear. Using the hospital narratives, cases were included in the sample if they included the words, “highheel,” “heels,” “high-heel,” “high heel,” “pumps,” “platform,” or “stiletto.” The NEISS provides stratified population weights which, with the *svy* function in Stata 17, were used to generate weighted estimates (with 95% confidence intervals) of the incidence of injury reports and, with Census estimates of the U.S. population [12], to produce population injury rates. Each case record also includes up to 2 diagnosis codes and a disposition. Diagnosis of fractures (versus sprain/strain, contusion/abrasion, or other diagnosis), and case disposition of held for observation, transferred, or admitted (versus treated and/or examined and released) are used to assess the severity of reported injuries. Cases are coded as fractures if either primary or secondary diagnosis is listed as a fracture.

To compare the pandemic period to the pre-pandemic period, the date cutoff of March 15, 2020 was used – the date the Centers for Disease Control and Prevention records as when U.S. states began to shut down to prevent the spread of COVID-19 [13]. The frequency and characteristics of HHSRIs before and after that date is used to assess a possible pandemic effect.

## Results

Based on 2,485 individual reports in the NEISS database with narrative text suggesting high-heel related injuries, there were an estimated 70,290 such injuries reported at EDs from 2016 through 2020 (95% confidence interval: 52,001 to 86,985), or an annual average of 14,058. However, only 6,290 of those injuries were reported in 2020 (CI: 4595 to 7985), marking a sharp drop from the average of 16,000 in the years 2016 to 2019. Fig. 1 (panel A) shows there was a 60-day average of 20 to 75 estimated visits per day from 2016 through early 2020, with seasonal peaks in the spring and fall (except for the spring of 2019). In 2020, however, there was a sharp drop in daily cases after March 15, as seen in panel B, which shows the slope of the cumulative trend.

**Fig. 1.**
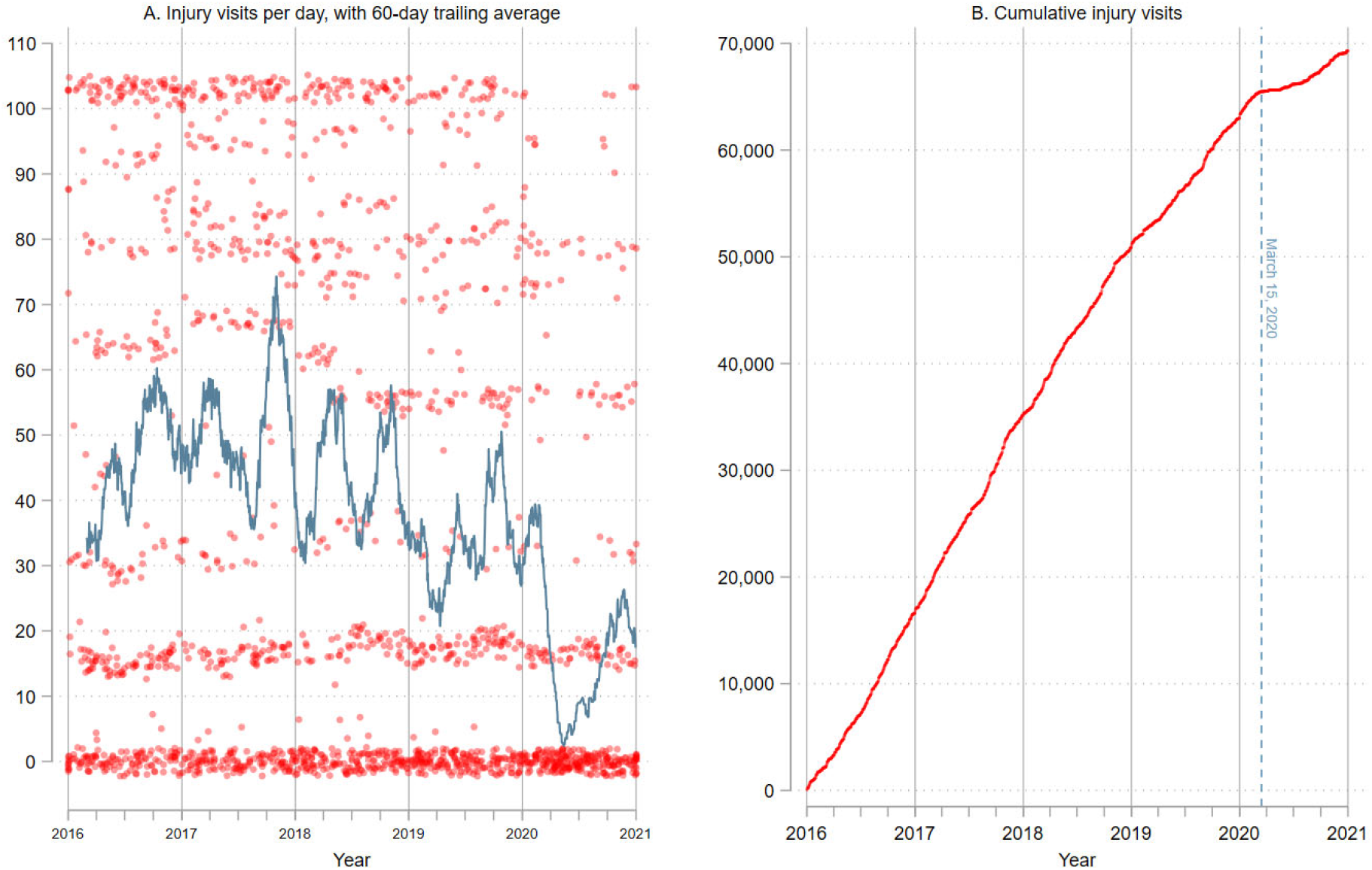
High-heel injury emergency department visits for U.S. women ages 15-69: 2016-2020”, Note: Data in panel A is top-coded at 103 (the top 10%), and markers jittered, for presentation.

There were 5.40 HHSRIs per 100,000 women ages 15-69 in 2020 (CI: 3.95 to 6.86), significantly below the peak of 16.41 in 2017 (CI: 12.09 to 19.86) (Fig. 2, panel A). Panel B of Fig. 2 compares the period from January 1 to March 15 in each year, and finds no significant change from 2016 to 2020, while panel C reveals the drop in 2020 relative to previous years occurred after March 15 – the start date of states shutting down in response to the COVID-19 outbreak.

**Fig. 2.**
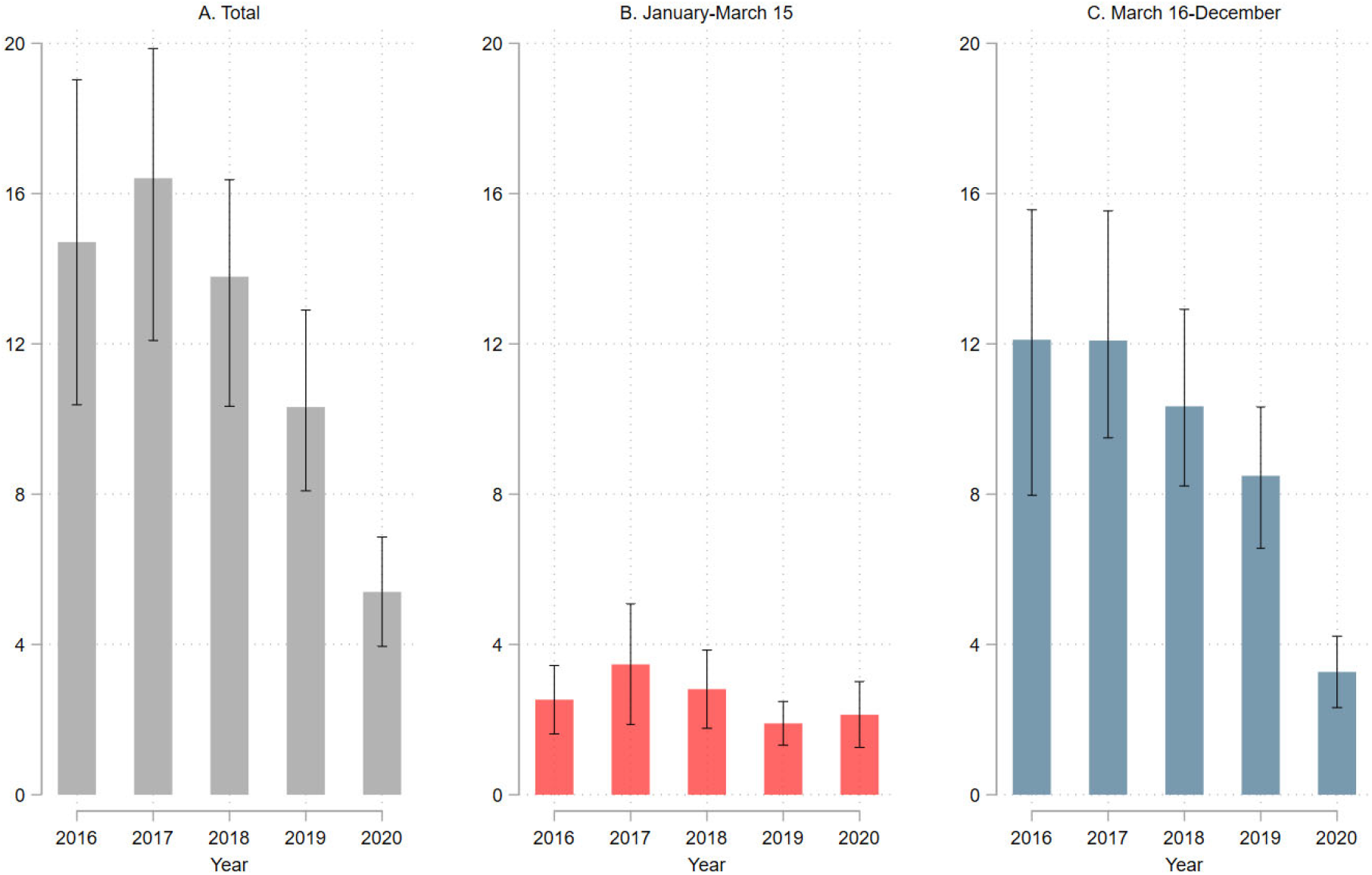
High-heeled shoe related injury visits per 100,000 women ages 15-69, 2016-2020 [95% CIs]

To assess whether the pandemic may have deterred women from reporting less severe injuries to EDs, the percentage of injuries diagnosed as fractures; and the percentage resulting in admission, transfer, or being held for observation, were calculated for the periods before and after March 15, 2020 (Table 1). Overall, 3.6% were admitted, transferred, or held for observation (CI: 2.4% to 5.3%), and 19.2% were diagnosed as fractures (CI: 16.5% to 22.2%), but there was no significant difference in either measure when periods were compared (Fig. 3). The pandemic period had an estimated 4.6% higher rate of fractures, but this was not significantly different from zero (*p* = .27); and a similarly insignificant 2.4% more cases admitted, transferred, or held for observation (*p* = .22).

**Table 1.** Diagnosis and disposition of high-heel shoe related injuries

| Diagnosis | % | LCL | UCL |
| --- | --- | --- | --- |
| Fracture | 19.2 | 16.5 | 22.2 |
| Sprain/strain | 43.9 | 40.3 | 47.6 |
| Contusion/abrasion | 8.7 | 6.6 | 11.4 |
| Other<sup>1</sup> | 28.2 | 24.4 | 32.4 |
| Disposition |  |  |  |
| Treated / examined and released<sup>2</sup> | 96.4 | 94.7 | 97.6 |
| Transferred, admitted, or held for observation | 3.6 | 2.4 | 5.3 |

<sup>1</sup> 21% are "other/not stated," other diagnoses are <3% each.
<sup>2</sup> Includes 0.5% left without being seen.
LCL, the lower 95% confidence limit of the estimate; UCL, the upper 95% confidence limit of the estimate.

**Fig. 3.**
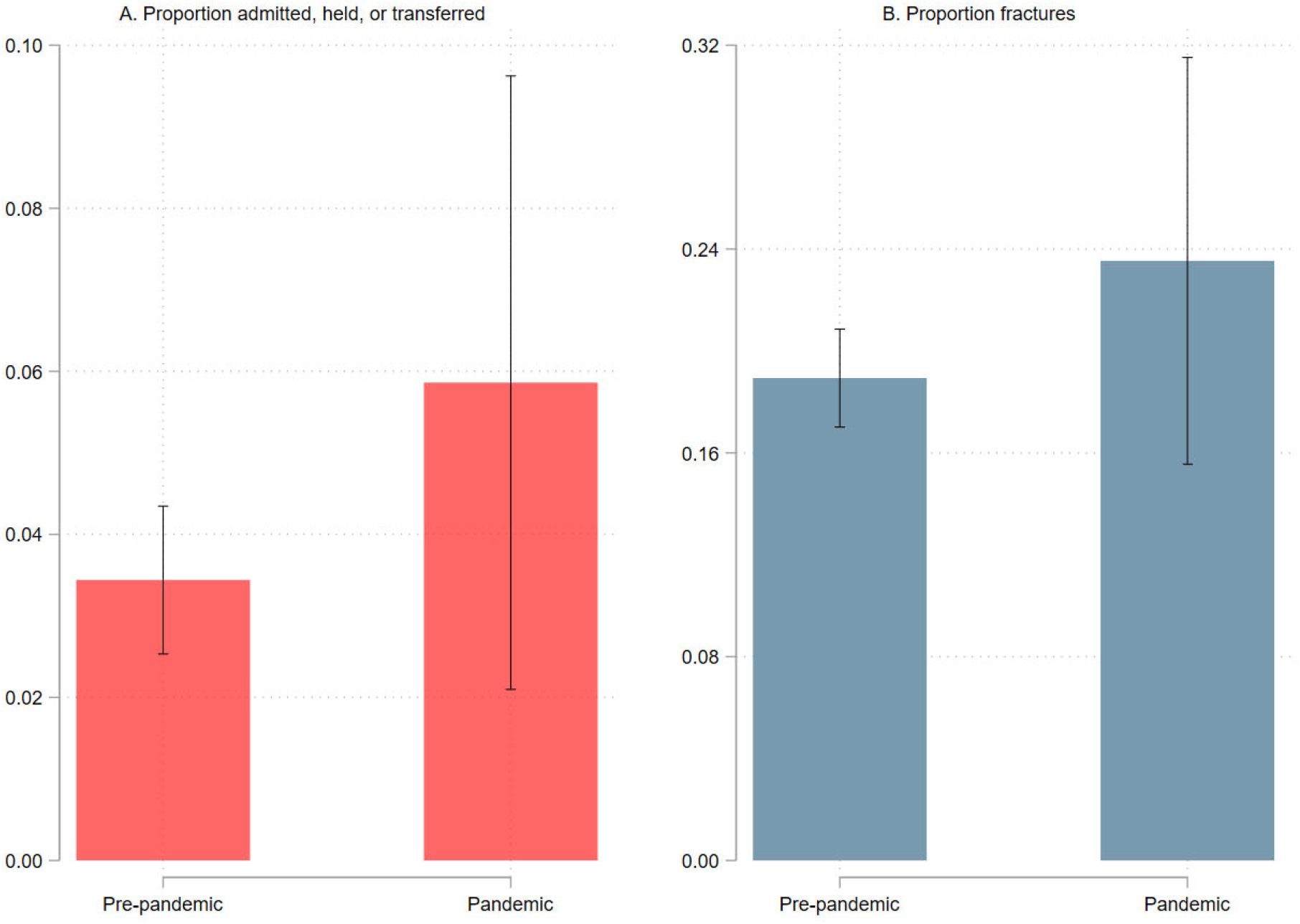
Characteristics of high-heel related injuries, pre-pandemic versus pandemic period [95% CIs]

## Discussion

Thousands of U.S. women are injured while wearing high-heeled shoes each year, according to reports from hospital EDs, as recorded in the NEISS. However, the onset of the COVID-19 pandemic – specifically, March 15, 2020 – was associated with a sharp decline in the rate of reported injuries related to high-heeled shoes among US women, as determined by product codes and text narratives. Analysis of case diagnoses and dispositions showed that more serious injuries – those coded as fractures, and those resulting in hospital admission, transfer, or observation – were estimated to be more common in the pandemic period, which might be the case if women with more minor injuries were staying away from hospitals, but these differences were not statistically significant.

These results are consistent with the possibility that reduced socializing, and more working from home, reduced the tendency of women to wear high-heeled shoes, thus decreasing their chance of injury. Some analysts suggest that pandemic adaptations, such as working from home, will result in new habits and permanent changes to common practices [14]. If that also is the case with regard to women’s footwear, the result may be a long term reduction in injuries caused by wearing high heels.

## Data Availability

All data produced are available online at: https://osf.io/t57gk/.

## Notes

### Competing Interest Statement

The authors have declared no competing interest.

### Funding Statement

This study did not receive any funding.

### Author Declarations

This study involves only openly available human data, which can be obtained from: http://www.cpsc.gov/library/neiss.html

